# Prognostic early snapshot stratification of autism based on adaptive functioning

**DOI:** 10.1101/2022.08.01.22278267

**Authors:** Veronica Mandelli, Isotta Landi, Elena Maria Busuoli, Eric Courchesne, Karen Pierce, Michael V. Lombardo

## Abstract

A major goal of precision medicine is to predict prognosis based on individualized information at the earliest possible points in development. Using early snapshots of adaptive functioning and unsupervised data-driven discovery methods, we uncover highly stable early autism subtypes that yield information relevant to later prognosis. Data from the National Institute of Mental Health Data Archive (NDA) (n=1,098) was used to uncover 3 early subtypes (<72 months) that generalize with 97% accuracy. Outcome data from NDA (n=2,561; mean age, 13 years) also reproducibly clusters into 3 subtypes with 99% generalization accuracy. Early snapshot subtypes predict developmental trajectories in non-verbal cognitive, language, and motor domains and are predictive of membership in different adaptive functioning outcome subtypes. Robust and prognosis-relevant subtyping of autism based on early snapshots of adaptive functioning may aid future clinical and research work (e.g., clinical trials, intervention), via prediction of these subtypes with our open web-based app (https://landiit.shinyapps.io/vineland_statification_proj/).

---

Autism is a clinical consensus label based on early difficulties in the domains of social-communication and restricted repetitive behaviors^1^. While the label of autism helps maximize consensus and reliability amongst clinical diagnostic judgments based on behavior, it may be less useful for many other important clinical and translational research objectives, such as honing in on differential biology, outcomes, and treatment responses^2,3^. With a view towards applying precision medicine^4,5^ to the field of autism, we should aim to move closer to labels that have higher utility for these types of objectives. As a step in this direction, a recent Lancet commission has proposed a call-to-action for more precise labels, such as ‘profound autism’, to identify the most profoundly affected individuals which require extra services and support^6^. Thus, a first-level distinction in the autism population should be made that separates out autistic individuals characterized by ‘disability’ versus ‘difference’ in developmental outcomes.

In this work, we characterize autism subtypes within the domain of adaptive functioning. There are several important reasons for why subtyping based on adaptive functioning may have immediate clinical and translational benefits. First, adaptive functioning is a pivotal domain with high ecological validity and predictive power for explaining later life outcomes (e.g., later independent living, educational attainment, employment)^7–10^ and also is associated with services and unmet needs^11^. Variability in adaptive functioning in the autism population is considerable, ranging from very profoundly affected individuals to those within typically-developing norms^12–14^.

Thus, distinctions within adaptive functioning are clearly needed to separate clinically-meaningful and outcome-sensitive heterogeneity in autism. Second, adaptive functioning can be quickly measured throughout the lifespan with standardized clinical assessment tools such as the Vineland Adaptive Behavior Scales (VABS). This offers potential for quick, repeated, and affordable assessments of an individual throughout the lifespan and can be deployed in multiple settings. Other advantages of the VABS are age-normalized scores and the ability to interpret minimal clinically significant change^12^. Third, changing adaptive functioning has become one of the key objectives for intervention research^12^. Thus, stratification models that can provide useful subtype labels may be important for facilitating advances in personalizing interventions. Fourth, adaptive functioning can be disentangled from an individual’s level of intellectual functioning^13^. While there is a relationship between adaptive functioning and large differences in intellectual functioning (e.g., contrasting individuals with IQ<70 versus IQ≥70), amongst those with IQ≥70 the variability in adaptive functioning is still considerable, ranging from highly affected individuals to individuals within the normative range for their age^12^. As individuals get older, the potential discrepancy between IQ and adaptive functioning can widen^13,15^. Thus, subtyping based on adaptive functioning may be able to capture real-world clinically-meaningful variability between individuals, even within the range of intact intellectual functioning.

Prior longitudinal work has attempted to identify subtypes based on differential trajectories of the VABS over the first two decades of life^14–17^. While this work is immensely important for describing how different types of individuals develop in terms of adaptive functioning, it cannot be utilized for early stratification in important clinical contexts such as intervention, because rich longitudinal information is not known about participants in such studies. A gap is present whereby there is a key need to be able to stratify individuals in developmentally/outcome-sensitive ways based on single snapshots of information at early stages in development. If there was a tool that allowed for highly robust and reproducible subtyping based on early single snapshots of adaptive functioning, this would potentially fill this gap and lead to further insights about how to predict treatment response and outcome in such individuals. Given such potential, we aimed to develop a stratification model that allows for data-driven discovery of robust and reproducible VABS subtypes based on a single snapshot of early VABS scores. We then show how subtypes are useful for predicting subsequent adaptive functioning outcome subtypes and developmental trajectories in non-verbal cognitive, language, and motor domains.

## Results

### Unsupervised data-driven identification of highly robust and reproducible autism adaptive functioning subtypes

In our first analysis we sought to test whether unsupervised data-driven stratifications could be made in autism based on early snapshots (<72 months) of adaptive functioning on the VABS. Our analysis approach applies stability-based relative clustering validation to identify data-driven clusters that are stable and reproducible in independent datasets (Fig 1). Here we find that a 3-cluster solution is unequivocally the best cluster solution that minimizes normalized cluster stability. This model produces very high generalization accuracy (97%) in independent data (e.g., the held-out NDA validation set) (Fig 2A; Supplementary Table 1). An equal male-bias is present in each of the 3 subtypes (*χ*^*2*^*(2)* = 3.64, *p* = 0.16). Importantly, the 3-subtype solution heavily deviates from the null hypothesis that the data originate from a single multivariate Gaussian distribution (*p* = 9.99e-5). Plots of the UMAP-reduced data shows evidence of two distinct peaks and a valley in between, where the 3rd subtype is located (Fig 2B). To better describe this 3-subtype solution, we next plotted VABS scores for each subtype and in each VABS domain (Fig 2C-D). Here we see a clear distinction between the subtypes in terms of ability, from high, medium, to low, that is preserved across each VABS domain. Although the subtype distributions overlap to some extent, the size of differences between subtypes is typically quite large (e.g., *Cohen’s d* > 1) for each pairwise subtype comparison (Fig. 2C-D) and these effect sizes are robustly preserved in independent training and validation sets.

**Figure 1:**
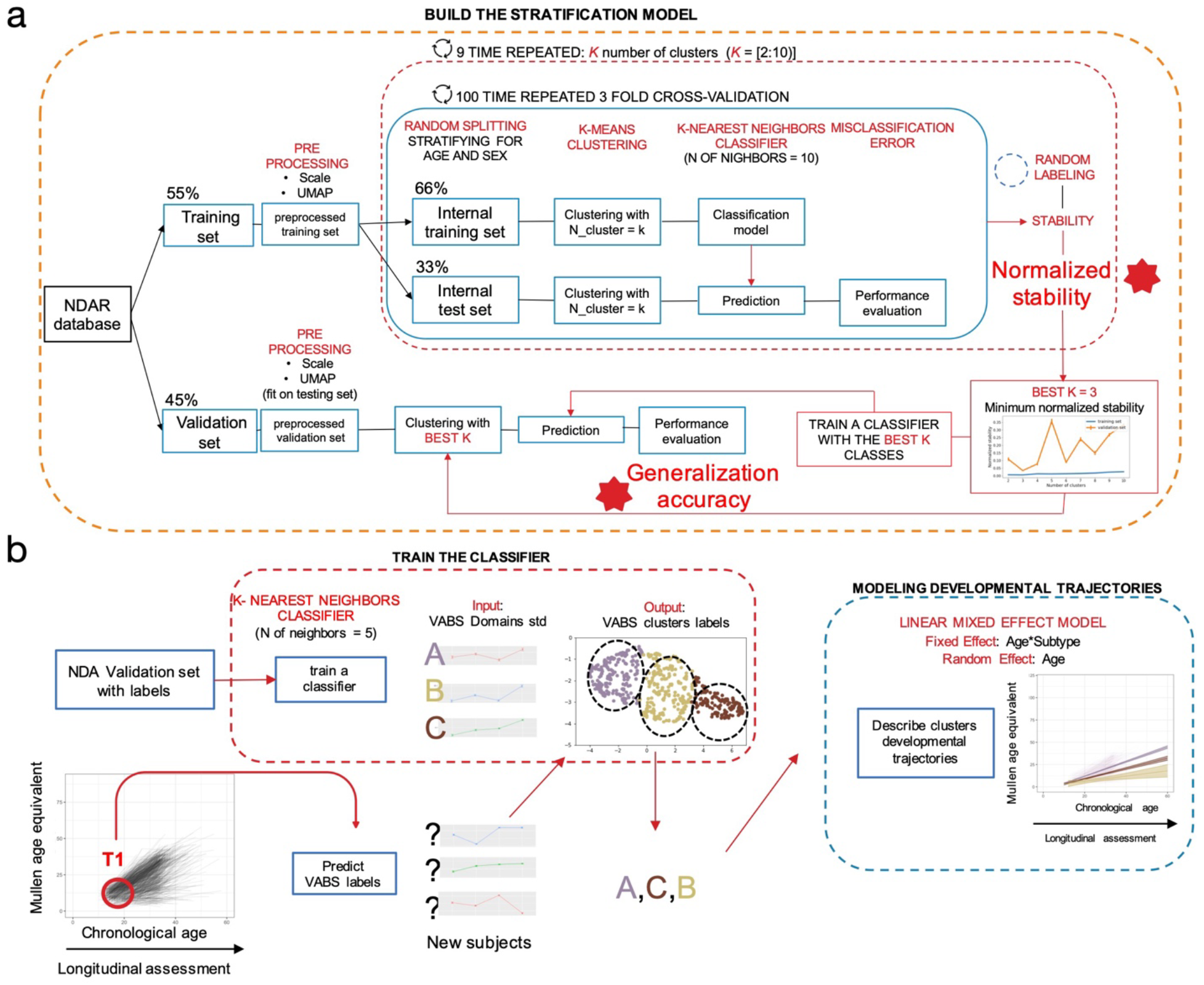
Schematic outlining the data analysis plan. The dataset used for initial data-driven discovery and validation through stability-based clustering is the NDA dataset (https://nda.nih.gov). NDA data is fed through the pipeline shown in panel A, which illustrates the reval algorithm pipeline. Once a robust and highly generalizable classifier is built from NDA, we apply that snapshot prediction model to the UCSD ACE longitudinal dataset. Panel B shows the analysis pipeline for applying the subtype prediction model and then modeling developmental trajectories.

**Figure 2:**
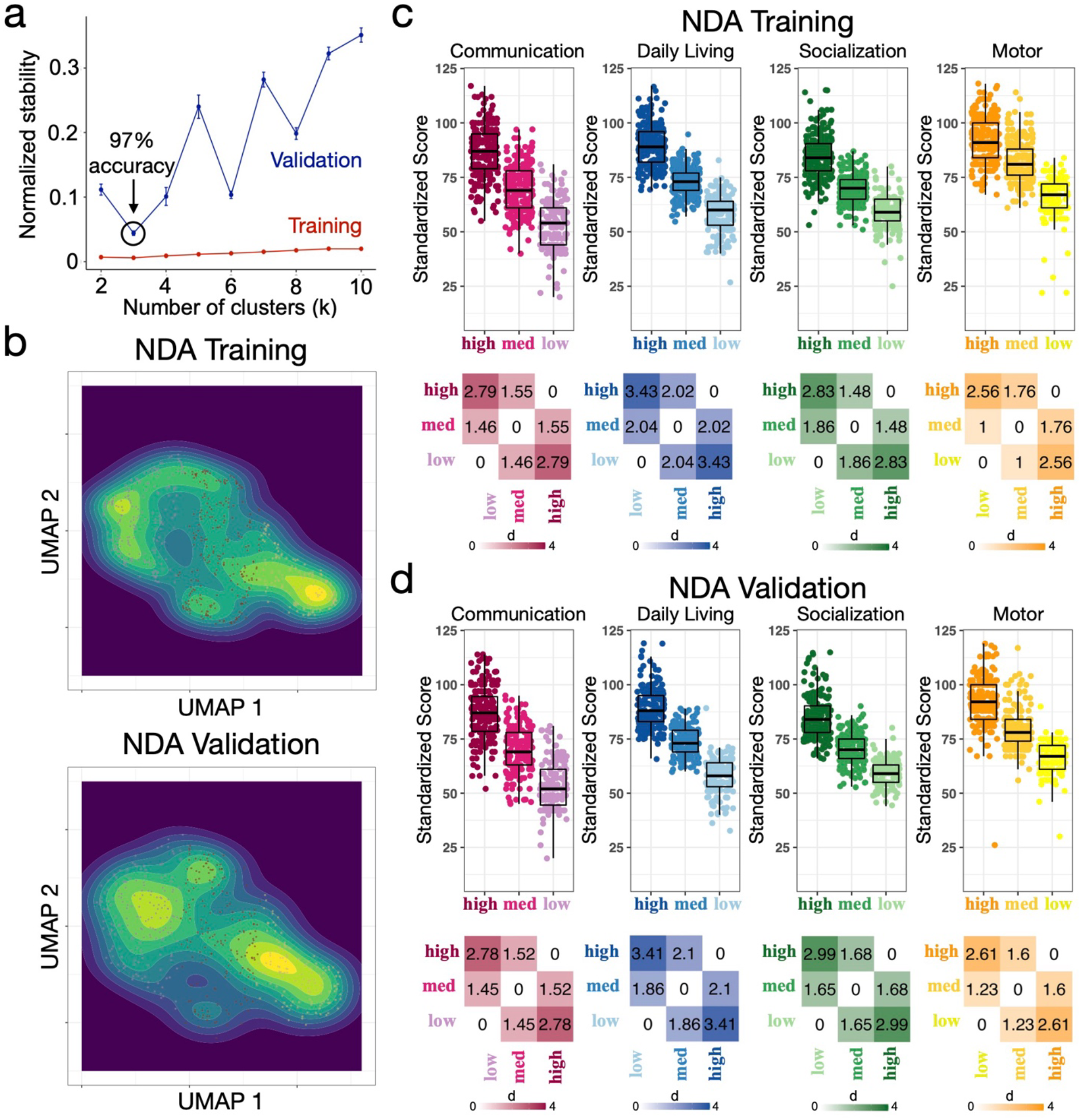
Unsupervised data-driven discovery of early snapshot autism adaptive functioning subtypes. This figure shows the results of stability-based relative clustering validation applied to the early snapshot NDA dataset. Panel a shows the normalized stability plot whereby k=3 is optimal solution minimizing normalized cluster stability. In an independent validation set, this k=3 solution generalizes with 97% accuracy. Panel b shows plots of UMAP-reduced data input into the clustering for both NDA Training (top) and Validation (bottom) sets. Density contours show the presence of two distinct peaks at either end of the UMAP 1 dimension and a valley in between. The individuals are colored by subtype (pink, green, blue) over the top of these density plots.Panels c-d graphically describe the subtypes across each VABS domain with scatter-boxplots and heatmaps depicting the pairwise standardized effect size difference between the subtypes. These plots are shown for both the NDA Training (panel c) and Validation (panel d) sets.

We next applied the same stratification approach to much older NDA outcome-relevant cohort (n=2,561; 6-61 years; mean age = 13 years). A 3-subtype solution emerges from this dataset with 99% generalization accuracy and strong rejection of the null hypothesis that data originate from a single multivariate Gaussian distribution (p = 9.99e-5) (Fig 3A-B). One of the subtypes can be considered an extreme outlier subtype since scores are at floor levels near 20 and with hardly any variability around this floor (Fig 3C-D). This subtype is relatively small in size (n=84) and comprises about 3% of all individuals in this older cohort. The remaining 2 subtypes can be described as relatively high or low and are both relatively large and equal in size (high n=1,313; low n=1,164).

**Figure 3:**
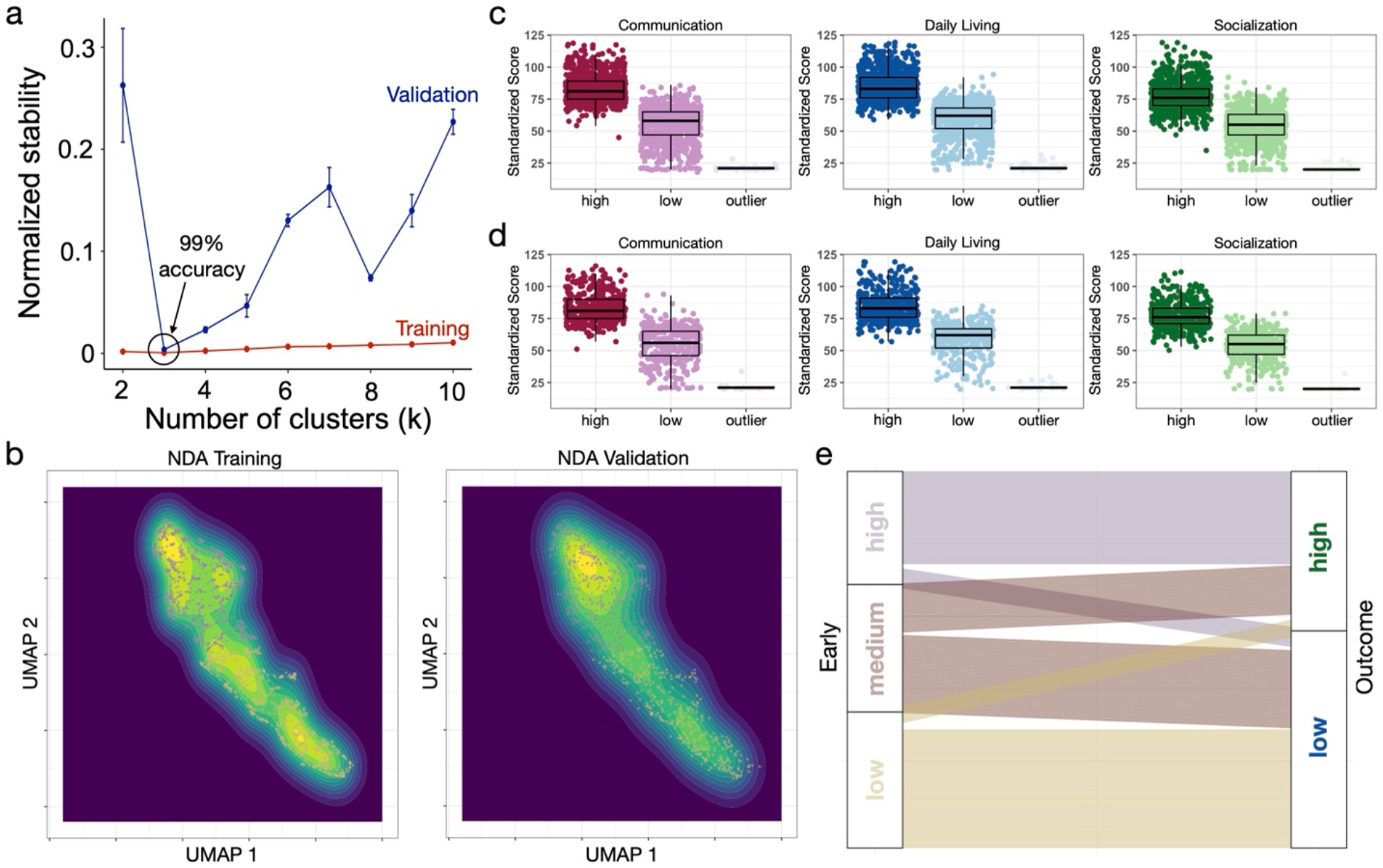
Later life outcome stratification. Panel a shows the normalized stability plot, indicating that the best clustering solution is k=3. This solution achieves 99% accuracy when generalizing to the validation set. Panel b shows UMAP scatter and density plots showing the high and low subtypes. Panels c (NDA Training) and d (NDA Validation) describes the subtypes for each of the 3 VABS scales (Communication, Daily Living, and Socialization). Panel e shows an alluvial plot of the n=126 subset of individuals that were in both early snapshot (left) and later life outcome (right) cohorts. Around 87% of the early snapshot high and low individuals stay in outcome high or low subtypes, whereas the early snapshot medium subtype splits between 41% in the high outcome subtype and 59% in the low outcome subtype.

### High and low early snapshot subtypes remain high and low at outcome

While early development is quite variable, later life outcomes tend to be much more stable^6,16^. Studies examining predictors of later-life adaptive functioning have shown that better adaptive functioning earlier in life predicts better later-life outcomes^14^. Thus, the presence of a high and low group at early ages may be prognostically suggestive retained good versus poor later-life outcome. However, the presence of a third subtype in between these two (e.g., the medium subtype) may suggest that this subtype is more uncertain regarding their later-life outcomes. To test these predictions, we examined correspondence of subtype labels in a subset of n=126 individuals present in both the NDA early snapshot and older outcome datasets. As predicted, individuals in either early high or low subtypes are highly probable of remaining in that same high or low subtype at outcome (high = 86%, 95% CI = 75-96%; low = 88%, 95% CI = 78-97%; high and low combined = 87%, 95% CI = 80-93%). In contrast, the early snapshot medium subtype is much more ambiguous with respect to later subtype outcomes, with about 41% (95% CI = 24-57%) moving to the high outcome subtype, while the remaining 59% (95% CI = 42-75%) move to the low outcome subtype (Fig 3E). Corroborating this finding as well as prior work^14^, longitudinal VABS data show relatively flat or slightly declining standardized score group-level trajectories throughout the time period up to 72 months and with no differences in trajectories between the subtypes (Supplementary Figure 1; Supplementary Tables 2-3).

### Adaptive functioning subtypes are sensitive to differential trajectories in non-verbal cognitive, language, and fine motor domains

In our next set of analyses we looked to examine whether autism early adaptive functioning subtypes are distinctions that are sensitive to differential trajectories in non-verbal cognitive, language, and fine motor domains, as measured by the MSEL up through the first 5 and a half years of life. In longitudinal NDA data, we find age*subtype interactions throughout MSEL VR, EL, RL, and FM subscales. These differences in developmental trajectories are most pronounced for the low subtype compared to high or medium subtypes, while differences between the high and medium subtype are less strong (Fig. 4A) (Supplementary Tables 2-3). Because clustering was applied to the first timepoint of NDA data, these results are not independent of the clustering procedure and may be biased. Thus, in the next analysis we tested for subtype differences in MSEL developmental trajectories using a large and completely independent dataset – the UCSD ACE dataset (n=1,216). Again, we discover age*subtype interactions across all MSEL subscales. These effects are driven by significant age*subtype interactions for all pairwise between-subtype comparisons except for the fine motor subscale, whereby strong differences were much less apparent (Fig. 4B) (Supplementary Tables 2-3). Including sex as covariate in our longitudinal models resulted in nearly identical results (Supplementary Tables 2). These results provide strong independent replication of the subtype MSEL trajectory differences. Overall, these results indicate that autism adaptive functioning subtypes isolated from early snapshots from the VABS are predictive of later trajectory differences across a range of developmental domains such as non-verbal cognitive ability, language, and fine motor skills.

**Figure 4:**
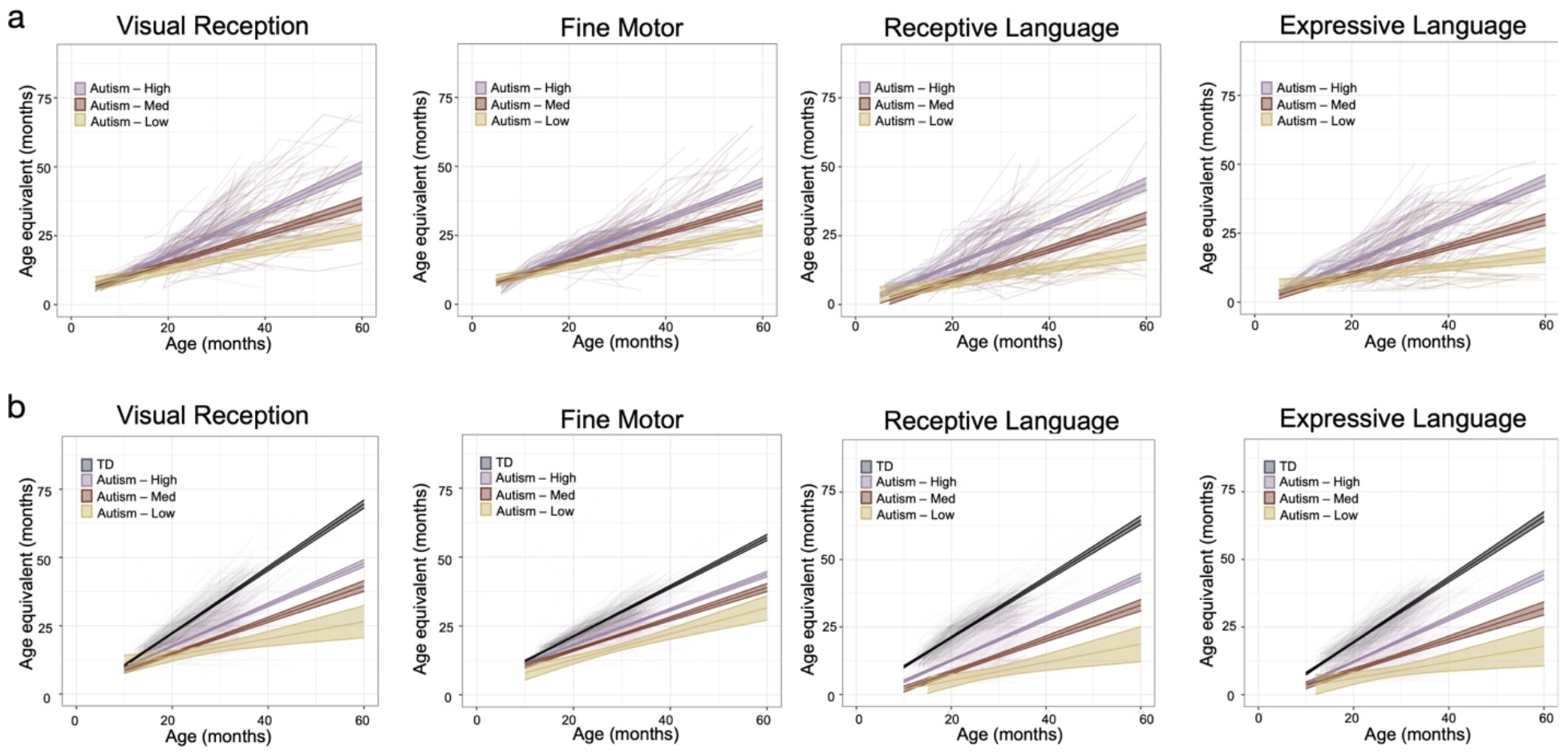
Differences in developmental trajectories between autism adaptive functioning subtypes in non-verbal cognitive, language, and fine motor domains measured by the MSEL. In both panels (NDA in panel A; UCSD ACE in panel B), we show spaghetti plots of the developmental trajectories for MSEL Visual Reception (VR), Expressive and Receptive Language (EL; RL) and Fine Motor (FM) subscales. Individual trajectories are shown in the background as transparent lines, while subtype trajectories (light purple, high; maroon, medium; cream, low; black, TD) are shown in solid lines with 95% confidence bands around them.

### Comparing and combining the autism adaptive functioning subtype model with normative models

Our clustering analysis showcase evidence that autism is not one homogeneous population with respect to early or later-life adaptive functioning. We have also showed that early snapshot adaptive functioning subtypes are outcome-relevant and developmentally-sensitive to variability in non-verbal cognitive, language, and motor trajectories. However, is the early snapshot subtyping model better than other competing models at explaining developmental trajectories? To answer this question we compare the early snapshot subtyping model to a normative model that uses typical-development defined age-standardized norm cutoffs for adaptive functioning on the VABS (VABS norms). The VABS norms model uses 1 and 2 SD cutoffs below the mean to create 3 subtypes. These subtype labels are then used in longitudinal MSEL models of the UCSD ACE data (Supplementary Tables 2-3). Model comparison AIC statistics were computed for both the *reval* subtype and VABS norm models. We find that the VABS norm model produces lower AIC values than the *reval* subtype model and with ΔΑΙC values greater than 10. This indicates that traditionally-defined ‘disability’ subtypes from VABS norms predict developmental trajectories as well or better than the data-driven *reval* autism subtype model on its own. We next created a ‘hybrid’ model, based on the combination of the *reval* autism subtype and VABS normative model labels (Fig 5A-B) (Supplementary Tables 2-3). This hybrid model produces the lowest AIC values and are indicative of a much better model than either *reval* subtyping or the VABS norm model alone (ΔAIC > 10) (Fig. 5C). Thus, improved utility for predicting cognitive and motor developmental trajectories could be facilitated through the combination of both a data-driven autism-specific subtyping approach combined with information regarding where the child stands relative to typically-developing adaptive functioning norms.

**Figure 5:**
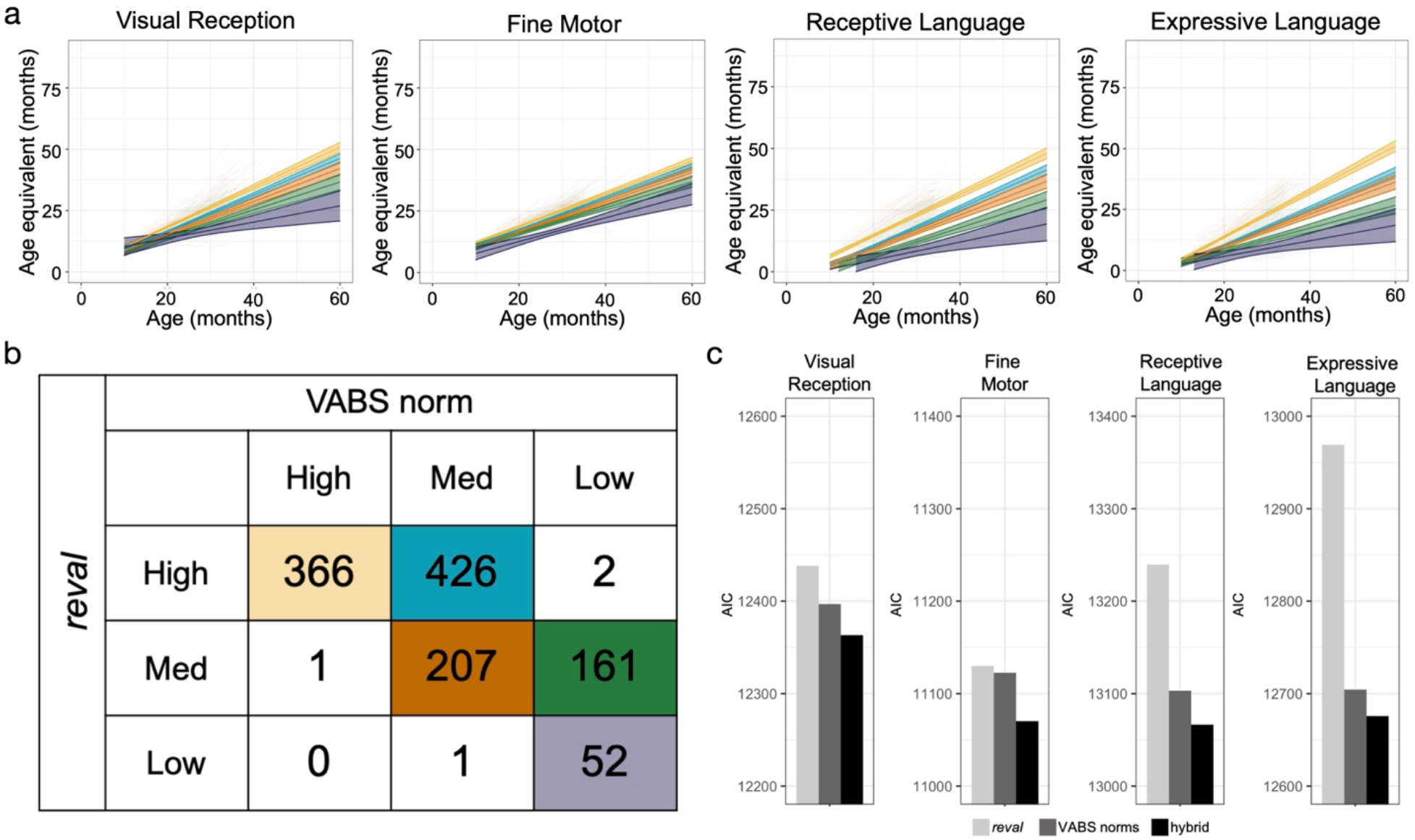
Model comparison between the autism-specific reval subtyping, VABS normative model, and a hybrid reval+VABS norms model. The hybrid model was built combining the adaptive functioning subtypes labels obtained using reval with those obtained by stratifying using the VABS norms. Panel A shows spaghetti plots of the developmental trajectories for MSEL Visual Reception (VR), Expressive and Receptive Language (EL; RL) and Fine Motor (FM) subscales for the 5 hybrid subtypes. The subtype colors are indicative of the subtypes shown in the cells of the confusion matrix in panel B. This confusion matrix indicates where individuals fall with respect to either the reval or VABS norm subtyping models. Panel c shows Akaike Information Criteria (AIC) model comparison statistics for comparing the reval (light gray), VABS norm (dark gray), and hybrid (black) models.

Finally, in order to aid future use in clinical or research settings, we developed a web application that will allow users to input VABS scores and receive subtype labels as output (https://landiit.shinyapps.io/vineland_statification_proj/). The intent behind this tool is to allow the field to immediately begin using these subtype labels for a priori experimental design in future studies. Furthermore, at an individualized level, the app may be useful for giving expectations behind developmental progress of an individual, given their subtype. For example, using the beta coefficients from the developmental trajectory models, we can express expected rate of growth per month in terms of age-equivalent scores for each domain on the MSEL (Supplementary Table 4).

## Discussion

In this study we aimed to identify developmentally and outcome-sensitive stratifications of the early autism spectrum based on adaptive functioning profiles that can be estimated via a simple, easy to administer, and well-known test of adaptive functioning in psychopathology - the VABS. Although the spread in scores on adaptive functioning measures like the VABS seem continuously distributed, the presence of robust and stable clusters indicates that autism is not one homogeneous population with respect to adaptive functioning. We show that in early development, there are 3 subtypes that can be described in terms of high, medium, and low ability. At older outcome ages (e.g., mean age 13 years old) autism can also be robustly clustered into 3 subtypes. One of these subtypes is an extreme outlier with very poor scores. The other two subtypes correspond to high versus low ability subtypes. We also discovered that some of the early snapshot subtypes hold information sensitive to later outcome. Both early high and low subtypes are highly likely to remain in those same subtypes at later ages. The early medium subtype is more uncertain in terms of later outcome subtype membership. This result indicates that early subtype stratification is possible and the resulting information can be outcome-sensitive, especially if individuals fall into the high or low subtype. For individuals falling into the early medium subtype, it may be pertinent for future work to examine what might be the predictors that nudge these individuals into better or worse outcome subtypes (e.g., influence of intervention, comorbid ADHD, executive functioning, environmental or educational influences, etc)^15,18–20^. In either situation (early high or low versus medium) the information can be used in highly clinically-useful ways and be important for designed future research studies (e.g., a priori stratification in clinical trials).

The discovered subtypes also hold predictive information about trajectories outside of adaptive functioning, such as non-verbal cognitive, language, and motor domains measured by the MSEL. Evidence of age*subtype interactions suggest that the subtypes show significant differences in the steepness of their slopes over age (e.g., rate of growth), with the most accelerated development present in the high group, while the more incremental and slowest growth is exhibited by the low group. This result suggests that our subtyping approach yields robust and reproducible subtype labels that are developmentally-sensitive outside of adaptive functioning and which could be utilized to inform expectations about prognosis in these domains. Finally, while the subtype model explains significant variance in developmental trajectories, the model could be further refined by simple knowledge of where an individual stands with respect to TD adaptive functioning norms. Combining the knowledge of both TD-defined norms and autism-specific norms for subtyping, we constructed a hybrid model, where labels are informed by where an individual is relative to both TD and autism norms. This hybrid model was proven to be the best of all models at predicting variance in non-verbal cognitive, language, and motor trajectories. Therefore, one key utility of our data-driven autism-specific subtyping approach is that it provides useful labels that when combined with VABS norms about typical development, can significantly enhance our precision at estimating expected growth for autistic individuals in each subtype.

It is important to underscore that our clustering approach allows us to not only identify the optimal number of clusters, but also describes how stable and generalizable that solution is to unseen new datasets. This 3-cluster early snapshot model is 97% accurate, while the outcome subtype model is 99% accurate. Given this high level of generalization accuracy combined with the breadth of the dataset in which it was defined (e.g., NDA data; n>1000), this result allows for high levels of confidence that these are stable and highly robust subtypes present in the autism population. This high level of confidence in generalization allows for immediate application of this subtyping model in new clinical and research work. This immediate impact and ability to re-use our stratification model is novel with respect to most studies using unsupervised data-driven techniques. A large limitation of most studies using traditional clustering techniques is that those studies are descriptive of what occurs in the dataset in question, but gives no indication about the generalizability, nor does it provide an easy-to-use stratification tool for the field to immediate apply in future clinical or research contexts. Our approach immediately translates unsupervised data-driven discoveries into supervised knowledge that can be used in future studies to significantly further progress in the field. Furthermore, a unique facet of our work is based on the idea of identifying developmentally/outcome-sensitive subtype information from early single snapshots of adaptive functioning in autistic individuals. A primary high-value application of such an approach would be in a context like treatment research. However, it should be noted that this early snapshot approach is distinct from the type of information gleaned from other work that tries to subtype based on differential trajectories on the VABS scores over the first two decades of life^14– 17^. The latter attempts to utilize known trajectories to derive subtypes. While such models are fit for optimally explaining within-individual developmental change, the opportunity for using it to aid early developmental research and clinical contexts (e.g., planning intervention) is limited. In contrast, our approach attempts to identify future predictive information via subtypes derived from single snapshots of VABS scores at early ages. Thus, the clinical and research applications for this niche are considerable and somewhat different from the other approaches that are more fit to describing subtypes based on observed trajectories.

In conclusion, we have demonstrated that a single snapshot of early adaptive functioning from the VABS can be utilized to predict highly robust and reproducible data-driven subtype labels that are informative about differential outcomes in adaptive functioning as well as different developmental trajectories in areas like non-verbal cognitive ability, language, and motor behavior. Because our clustering approach allows for immediate translation of unsupervised data-driven discoveries into supervised knowledge (e.g., a classifier), this work stands out in being able to allow others in the field to directly utilize our subtyping model in their own future work. Our open web-based tool (https://landiit.shinyapps.io/vineland_statification_proj/) allows individuals to insert their own VABS data and get subtype labels and developmental predictions about such subtypes as output. We hope that this work enables future research discoveries and is useful to help clinicians get initial expectations about prognosis based on limited early snapshots of adaptive functioning that are likely to be routine parts of initial clinical assessments.

## Methods NDA dataset

To first identify robust and reproducible early snapshot subtypes using unsupervised data-driven methods, we utilized large publicly available phenotypic data from the National Institute of Mental Health Data Archive (NDA). Querying NDA in March 2020, we identified children with a diagnosis of autism ages 6-72 months with at least one set of either Vineland Adaptive Behavior Scales (VABS) II parent and caregiver form, VABS-II survey form, or VABS-3^21^ (Fig. 1A). After data merging and cleaning, n=1,098 individuals from n=48 different originating datasets remained for further downstream analysis. A subset of these individuals also had longitudinal VABS data (n=410). For the cross-sectional early snapshot clustering analyses, only the earliest time-point was used from these individuals. However, in later follow-up analyses, we utilized the longitudinal subset for examining developmental trajectories. We also extracted another n=2,561 individuals from NDA with VABS scores at older ages (6-65 years). An identical data-driven clustering approach was used for this older ‘outcome’ cohort. See the Supplementary Methods for more details about the NDA dataset and Supplementary Table 5 for a list of NDA IDs and collections that these data come from.

### UCSD ACE dataset

A second independent dataset was also utilized, whereby we applied the NDA early snapshot subtype prediction model to predict subtype labels based on early VABS scores. This second dataset was longitudinal and allowed for an independent test of longitudinal hypotheses about the subtype model’s sensitivity to detect differences in developmental trajectories. This longitudinal dataset was collected at the University of California, San Diego Autism Center of Excellence (UCSD ACE) and comprises n=1,216 autistic children and n=689 typically-developing (TD) children, aged 9-72 months at their first Vineland assessment (T1) (Supplementary Table 6). UCSD ACE was sampled and ascertained through a combination of population-based autism risk screening at 12 months^22^ and community referrals. Longitudinal assessments took place approximately every 6 months from intake until outcome assessments around 4-5 years of age. Clinical diagnosis of autism was made at UCSD ACE by expert clinicians at the outcome assessment and was aided by a full battery of tests including VABS, the Mullen Scales of Early Learning (MSEL)^23^, and the Autism Diagnostic Observation Schedule (ADOS-2)^24^. These diagnoses have been shown to be highly stable even from very early ages^25^. Intake VABS scores from UCSD ACE were utilized to predict subtype labels from the NDA prediction model and then subsequent longitudinal modeling was implemented on the full set of UCSD ACE VABS or MSEL scores to test for trajectory differences. For more details about the UCSD ACE dataset please refer to the Supplementary Methods.

## Measures

### Vineland Adaptive Behavior Scales (VABS)

The VABS is a widely used standardized and semi-structured parent interview for assessing adaptive functioning in typical and clinical developmental populations throughout the lifespan. For children <72 months, the VABS assesses adaptive functioning in four major domains: communication, daily living skills, socialization, and motor skills. For individuals beyond 72 months, only the communication, daily living skills, and socialization domains are used. Standardized scores for each domain are computed to indicate where an individual scores relative to typically-developing age-appropriate norms, whereby for each standardized score, the mean is 100 and the standard deviation is 15. For model comparison purposes, we utilized these typically-developing norms to construct VABS normative subtypes, using cutoffs of 1 and 2 standard deviations below the mean.

### Mullen Scales of Early Learning (MSEL)

*The MSEL* is a standardized developmental test that can be administered from birth to 68 months of age and assesses development of non-verbal cognitive, language, and motor skills. Four of the five MSEL subscales were utilized in this work: Visual Reception (VR), Expressive Language (EL), Receptive Language (RL), Fine Motor (FM). In this work, we examined growth over time in age equivalent scores for each of these subscales. MSEL data was available for both NDA and UCSD ACE datasets. For evaluating MSEL developmental trajectories, we utilized MSEL age-equivalent scores.

## Statistical Analyses

### Stability-based relative clustering validation

Unsupervised data-driven clustering was achieved using our Python library (*reval*; https://github.com/IIT-LAND/reval_clustering) for implementing stability-based relative clustering validation^26,27^ (see Supplementary Methods for more details) (Fig 1). To reproduce this analysis please see our code deposited here: https://github.com/IIT-LAND/vineland_subtyping. Our choice of clustering and classification algorithms to utilize throughout application of *reval* was k-means clustering and k-Nearest Neighbors classification. For the NDA early snapshot dataset (6-72 months), we used a 55-45 training-validation split, while ensuring age and sex were balanced across this split. Due to the much larger sample size of the older NDA outcome dataset, we used a 67-33 training-validation split. After data are split into training and validation sets, we implemented preprocessing steps such as imputing missing values with a k-Nearest Neighbors imputation algorithm (*sklearn*.*impute*.*KNNImputer*), scaled data to a mean of 0 and standard deviation of 1 (*sklearn*.*preprocessing*.*StandardScaler*), and then applied Uniform Manifold Approximation and Projection (UMAP)^28^ dimensionality reduction (n_neighbors = 30, min_dist = 0.0, n_components = 2, random_state = 42, metric = Euclidean). All preprocessing steps fitted on the training set, were then applied to the validation set. After preprocessing, we apply a 3-fold internal cross-validation scheme to identify the optimal number of clusters (k-range from 2-10) that minimizes normalized cluster stability. This internal cross-validation scheme was repeated 100 times with different cross-validation splits to ensure robustness. Finally, a grid search procedure was utilized to select optimal hyperparameters (e.g., k=5 neighbors for the k-Nearest Neighbor classifier) utilized throughout. The final optimal k identified from the training set was applied to the validation set for k-means clustering, and then a k=5 k-Nearest Neighbor classifier was fit to the training set and then applied to predict clustering labels on the validation set. The generalization accuracy on the validation set is then computed by comparing the classifier’s predicted labels to the actual clustering labels identified in the validation set.

While stability-based relative clustering validation in *reval* tells us about the stability of clustering solutions, it does not test whether the actual solution is indicative of true clusters. Therefore, we followed up on the *reval* analysis by using the *sigclust* library in R to test whether the observed clustering solution significantly differs from the null hypothesis that the data originates from a single multivariate Gaussian distribution^29^.

### Longitudinal data analyses

To test for subtype differences in developmental trajectories, we utilized longitudinal data present in NDA and a second independent dataset - UCSD ACE. After *reval* identified the optimal clustering solution that generates high levels of generalization accuracy, we built a prediction model (i.e. k-nearest neighbors classifier) that learns the subtype distinctions from NDA data and applied this model to both NDA and UCSD ACE longitudinal datasets. For these prediction models we utilized the earliest VABS observation per each longitudinal subject, after identical preprocessing steps as those described before implementation of *reval* (e.g., imputation, scaling, UMAP dimensionality reduction). For testing developmental trajectory differences, we utilized linear mixed effect models (i.e. the *lmer* function in the *lme4* R library). The dependent variable in these models was either VABS domain standardized scores or MSEL subscale age equivalent scores. Fixed effects were specified as age, subtype, and the age*subtype interaction, while the random effect of age was modeled within-subject with random intercepts and slopes. Since multiple comparisons are made, we use FDR correction at a level of q<0.05. Follow-up tests for specific pairwise group comparisons were made to decompose and describe age*subtype interactions, and similar FDR control for multiple comparisons was used.

### Model comparisons

To evaluate the utility of the VABS early snapshot subtyping model against other types of models, we compared how well a model using VABS typically-developing norms (VABS norm; e.g., 1 and 2 SD cutoffs) could predict trajectories. Additionally, we also examined a *hybrid* model that combines *reval* subtype labels with the VABS norm model. Models were compared by evaluating how well they explain variance in longitudinal MSEL analyses, with the Akaike Information Criteria (AIC) being used as the model comparison statistic. Models with lower AIC scores are considered better and we also computed AIC difference scores (ΔΑΙC) as a quantitative indicator of just how much better the best model from the comparison model. ΔΑΙC>10 indicates that the comparison model has little to no support for being as good as the best model with the lowest AIC^30^.

## Supporting information

Supplementary Methods and Results

## Data Availability

All data utilized in this work can be found on the National Institute of Mental Health Data Archive (NDA; https://nda.nih.gov).

## Code availability

Analysis code to reproduce the analyses and figures can be found on GitHub (https://github.com/IIT-LAND/vineland_subtyping). The *reval* Python library can be found on GitHub (https://github.com/IIT-LAND/reval_clustering) and the documentation can be found here: https://reval.readthedocs.io. The Shiny app that allows users to input their own VABS data and get subtype predictions can be found here: https://landiit.shinyapps.io/vineland_statification_proj/.

## Acknowledgments

This project has received funding from the European Research Council (ERC) under the European Union’s Horizon 2020 research and innovation programme under grant agreement No 755816 (ERC Starting Grant to MVL). Data from UCSD ACE was collected with support from the following grants: NIMH R01-MH080134 (KP), NIMH R01-MH104446 (KP), NFAR grant (KP), NIMH Autism Center of Excellence grant P50-MH081755 (EC, KP), NIMH R01-MH036840 (EC), NIMH R01-MH110558 (EC), NIMH U01-MH108898 (EC), NIDCD R01-DC016385 (EC, KP, MVL).

## Author Contributions

Conceptualization: MVL, VM, IL. Methodology: MVL, IL, VM. Software: IL, VM. Formal analysis: MVL, VM, IL. Investigation: MVL, IL, VM, EMB, EC, KP. Data curation: MVL, IL, VM, EC, KP. Writing - original draft preparation: MVL, VM, IL, EMB, EC, KP. Writing - review and editing: MVL, VM, IL, EMB, EC, KP. Visualization: MVL, VM, IL. Supervision: MVL. Project administration: MVL. Funding acquisition: MVL, EC, KP.

