## Supplementary Methods and Results for "Prognostic early snapshot stratification of autism based on adaptive functioning"

### *Description of the NDA dataset*

The National Institute of Mental Health Data Archive (NDA) (https://nda.nih.gov) dataset was downloaded in January 2020 and is composed of subjects identified as having either the VABS-II Parent and Caregiver Rating Form, the VABS-II Survey Form, or the VABS-3. Available data originated from 48 different NDA collections. The subjects included in this study are indicated by their NDAR GUIDs along with their collection IDs reported in Supplementary Table 5. All VABS data were combined and then data was filtered to only include individuals with an autism diagnosis and VABS data collected between 6-72 months of age. Duplicate data were identified and dropped. For individuals with repeated measures VABS data available (i.e. longitudinal data), for the clustering analyses, we used the earliest of these repeated measures VABS data. Finally if more than one VABS subscale domain was missing, the subject was dropped from the analysis. For individuals missing only one VABS subscale domain, we utilized k-nearest neighbor imputation to impute the missing score.

### *Description of the UCSD ACE dataset*

Description of the UCSD ACE dataset can be found elsewhere in previously published work using this dataset[1–4]. Here we briefly describe the dataset and what data was used for the current study. Toddlers were recruited through two mechanisms: community referrals (e.g., website) or a general population-based screening method called *Get SET Early*[5] that allowed for the prospective study of ASD beginning at 12 months based on a toddler's failure of the CSBS-DP Infant-Toddler Checklist[6]. All toddlers were tracked from an intake assessment around 12 months and followed roughly every 6 months until 3–4 years of age. All toddlers, including normal control subjects, participated in a series of tests collected longitudinally across all visits, including the Autism Diagnostic Observation Schedule (ADOS), MSEL, and VABS. Only VABS and MSEL data is utilized in the current study and all available longitudinal data across these two measures were used. All testing occurred at the University of California, San Diego Autism Center of Excellence (ACE). Work utilizing this dataset was approved by the Institutional Review Board at University of California, San Diego. Parents provided written informed consent according to the Declaration of Helsinki and were paid for their participation. As of March 2020, n=1,216 autistic children were assessed with the VABS at least once before 72 months, and n=1,199 were also assessed with the MSEL. See Supplementary Table 6 for a breakdown of sample sizes and numbers of males and females at each timepoint.

### *Stability-based relative clustering validation*

Unsupervised data-driven discovery methods such as clustering were used within a stability-based relative clustering validation framework[7,8]. The aim of our clustering validation approach was to identify autism adaptive behavior subtypes based on data-driven patterning across VABS subscales that are indeed representative of population-level stratification within the autism population. Our logic here is that if subtypes can be identified with unsupervised data-driven techniques such as clustering, we would strive to have such solutions that reveal stratifications of the population that would be present if we were to do similar kinds of clustering analyses on other independent datasets with the same VABS measures. Unlike more commonly used internal clustering approaches (e.g., silhouette technique), whereby the optimal number of clusters are learned within one dataset but where no evidence is provided to support generalizability or representativeness of such solutions in independent data, relative clustering validation approaches offer the unique ability to demonstrate that data-driven clustering solutions are indeed representative and generalizable in other independent datasets from the same population. By being able to demonstrate that data-driven solutions are indeed robust and reproducible in independent datasets, this proof-of-concept allows us to immediately translate such data-driven discoveries into supervised knowledge that could be used within the context of classification or prediction models applied to new datasets from the same population.

Our relative clustering validation analyses are implemented within our recently developed *reval* Python library[8] (https://github.com/IIT-LAND/reval_clustering). The *reval* algorithm is shown graphically in Fig. 1A and described in detail in Landi et al.,[8]. To briefly show how the *reval* algorithm works, *reval* begins by having users provide a training and test set. Within the training set the goal will be to estimate the optimal number of clusters. To achieve this goal *reval* implements an internal training-validation split on the original training dataset and then uses these internal training-validation sets to run through a range of cluster solutions implemented on each set. A classification model is then produced to learn the clusters in the internal training set and then used to identify those clusters in the validation set. From this an accuracy value will be extracted from the predictions (misclassification error). This procedure, combined with the accuracy values after random labeling, provides a performance metric, called stability, whereby lower values indicate the higher reproducibility of the solution. By inspecting the stability metric across a range of possible clustering solutions (k), the user can identify the optimal number of clusters as the solution (k) that minimizes stability. Once the optimal number of clusters (k) has been identified, we apply that k as the number of clusters to identify in the original held-out test set. A classification model is then trained based on the original training set and applied to predict the original held-out test set. This allows for accuracy to again be estimated between the actual cluster labels in the test set versus the classifiers predicted cluster labels. This accuracy estimated on the held-out test set is considered the generalization accuracy and allows the user to interpret whether the clustering solution identified is indeed robust and reproducible in independent datasets. It should be noted that throughout all clustering and classification models produced by *reval*, the user is free to select any number of possible clustering and classification techniques available within the *scikit-learn* python library, and thus does not force users to select a specific recipe of clustering and classification approaches. For more details about *reval* please see our prior published work[8].

Regarding how we applied *reval* to our existing datasets, we report here each step and the decisions that were made throughout (Fig 1B). To reproduce this analysis please see our code deposited here: https://github.com/IIT-LAND/vineland_subtyping. Our choice of clustering and classification algorithms to utilize throughout application of *reval* was k-means clustering and k-Nearest Neighbors classification. For the NDA early snapshot dataset (6-72 months), we used a 55-45 training-validation split, while ensuring age and sex were balanced across this split. Due to the much larger sample size of the older NDA outcome dataset, we used a 67-33 training-validation split. After data are split into training and validation sets, we implemented preprocessing steps such as imputing missing values with a k-Nearest Neighbors imputation algorithm (*sklearn.impute.KNNImputer*), scaled data to a mean of 0 and standard deviation of 1 (*sklearn.preprocessing.StandardScaler*), and then applied Uniform Manifold Approximation and Projection (UMAP)[9] dimensionality reduction (n_neighbors = 30, min_dist = 0.0, n_components = 2, random_state = 42, metric = Euclidean). All preprocessing steps fitted on the training set, were then applied to the validation set. After preprocessing, we apply a 3-fold internal cross-validation scheme to identify the optimal number of clusters (k-range from 2-10) that minimizes normalized cluster stability. This internal cross-validation scheme was repeated 100 times with different cross-validation splits to ensure robustness. Finally, a grid search procedure was utilized to select optimal hyperparameters (e.g., k=5 neighbors for the k-Nearest Neighbor classifier) utilized throughout. The final optimal k identified from the training set was applied to the validation set for k-means clustering, and then a k=5 k-Nearest Neighbor classifier was fit to the training set and then applied to predict clustering labels on the validation set. The generalization accuracy on the validation set is then computed by comparing the classifier's predicted labels to the actual clustering labels identified in the validation set.

***Supplementary Results***

***Longitudinal VABS trajectories***

Longitudinal VABS data was available in a subset of n=410 individuals within NDA. Analysis of the longitudinal data identifies relatively flat (e.g., Communication) or slightly declining trajectories (e.g., Daily Living, Socialization, Motor) over age for all subtypes (Supplementary Figure 1). Age-related decline is compatible with the known negative correlation between VABS scores and age[10]. Subtypes did not differ in trajectories, with the exception of the motor domain, whereby significant age*subtype interactions appear and are driven by declining trajectories in the high and medium subtypes, but a relatively stable and poor trajectory in the low subtype (Supplementary Figure 1) (Supplementary Tables 2-3). Similar longitudinal analysis of VABS data was applied to the UCSD ACE dataset. VABS subtype trajectories in UCSD ACE data are remarkably similar to those seen in NDA, with no differences between subtypes in trajectories and relatively flat group-level trajectory for Communication, and declining trajectories for all other domains (Supplementary Figure 1) (Supplementary Tables 2-3).

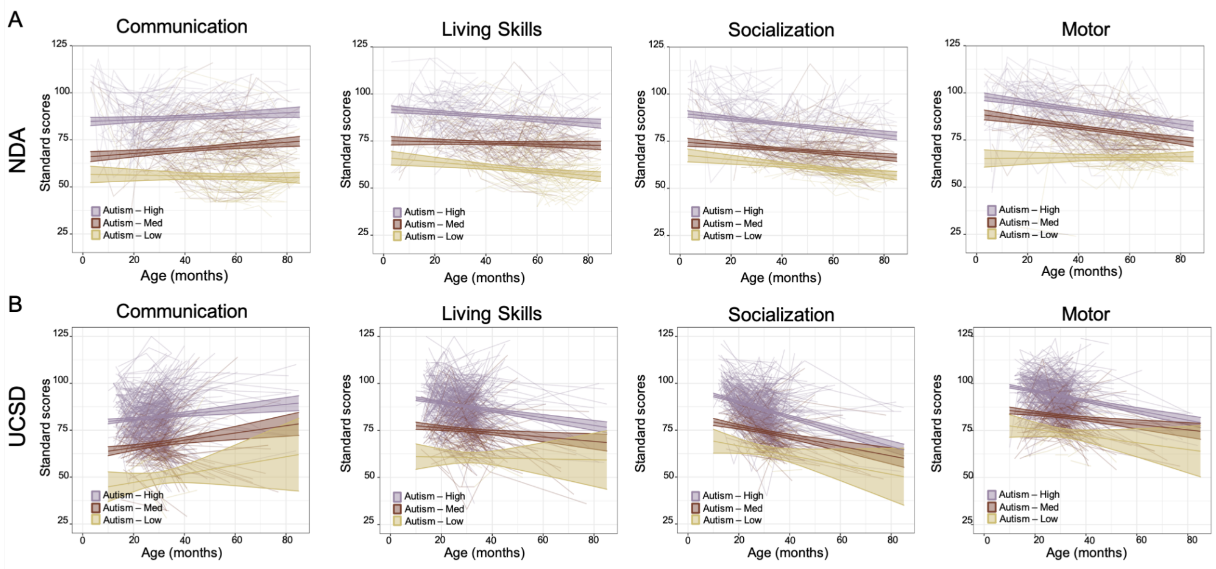


***Supplementary Figure 1: Adaptive behavior trajectories with longitudinal data from NDA (panel a) and UCSD ACE (panel b).*** *For each panel, we show trajectories for each domain within the VABS. Individual participants are shown with transparent lines, while group-level trajectories are shown as bold lines alongside 95% confidence bands.*

***Supplementary Table Legends***

***Supplementary Table 1***: Subtype sample sizes for the Early Snapshot and Outcome datasets.

***Supplementary Table 2***: Linear mixed effect model ANOVA tables and statistics for all longitudinal analyses.

***Supplementary Table 3*****:** Post-hoc pairwise group comparisons for longitudinal models.

***Supplementary Table 4*****:** Table indicating rate of growth per unit of age-equivalent (months) for each of the hybrid subtypes and for each MSEL subscale.

***Supplementary Table 5*****:** NDA subject IDs (subjectkey) and collection IDs for each subject from NDA in the Early Snapshot or Outcome datasets.

***Supplementary Table 6*****:** Descriptive statistics for the UCSD ACE dataset.
